# Subacute changes in brain functional network connectivity after nocturnal sodium oxybate intake are associated with anterior cingulate GABA/glutamate balance

**DOI:** 10.1101/2022.11.21.22282584

**Authors:** Francesco Bavato, Fabrizio Esposito, Dario A. Dornbierer, Niklaus Zölch, Boris B. Quednow, Philipp Staempfli, Hans-Peter Landolt, Erich Seifritz, Oliver G. Bosch

## Abstract

Sodium oxybate (γ-hydroxybutyrate, GHB) is an endogenous GHB/GABA_B_ receptor agonist, clinically used to promote slow-wave sleep and reduce next-day sleepiness in disorders such as narcolepsy and fibromyalgia. The neurobiological signature of these unique therapeutic effects remains elusive. Promising current neuropsychopharmacological approaches to understand the neural underpinnings of specific drug effects address cerebral resting-state functional connectivity (rsFC) patterns and neurometabolic alterations. Hence, we performed a placebo-controlled, double-blind, randomized, cross-over pharmacological magnetic resonance imaging study with a nocturnal administration of GHB, combined with magnetic resonance spectroscopy of GABA and glutamate (Glu) in the anterior cingulate cortex (ACC). Sixteen healthy male volunteers received 50mg/kg GHB p.o. or placebo at 02:30am to maximize deep sleep enhancement and multi-modal brain imaging was performed at 09:00am of the following morning. Independent component analysis of whole-brain rsFC revealed a significant increase of rsFC between the salience network (SN) and the right central executive network (rCEN) after GHB intake compared to placebo. This SN-rCEN coupling was significantly associated with changes in GABA and GABA/Glu levels in the ACC (p_all_<0.05). The proposed framework allows to identify a neural pattern of pharmacological modulation of the SN, which may serve as a neurobiological signature of the wake-promoting effects of GHB.

## Introduction

Sodium oxybate (γ-hydroxybutyrate, GHB) is an endogenous GHB/GABA_B_ receptor agonist, which regulates important homeostatic functions such as eating and sexual behavior, as well as the sleep-wake cycle [1]. The latter effect is clinically used in narcolepsy and fibromyalgia, where GHB strongly enhances nocturnal slow-wave sleep [2,3], while promoting next-day wakefulness [2,4]. Potential clinical applications of GHB to treat excessive daytime sleepiness have been suggested in different neuropsychiatric disorders including Parkinson disease and depression [5,6]. From a neurobiological perspective, these peculiar and complex clinical patterns are not yet fully understood.

Recent neuropsychopharmacological approaches involve the assessment of cerebral resting-state functional connectivity (rsFC) to understand how psychoactive substances differentially modulate brain functioning. Here, an established model to analyze rsFC on a large-scale level involves three core neurocognitive networks: the default mode network (DMN), the central executive network (CEN) and the salience network (SN) [7]. The DMN is located in the posterior cingulate cortex, the medial prefrontal cortex, and the inferior parietal lobule and has an active role for internally directed mental experiences [8]. The CEN is anchored in the dorsolateral prefrontal cortex and inferior parietal lobule and is mostly involved in goal-directed cognitive operations, such as decision making, problem solving and in actively maintaining and processing working memory [9]. Finally, the SN has a modulatory function in both rest and task-related activity. It responds to the degree of subjective salience of a stimulus, to control the activity of DMN and CEN, and thereby regulating the switch into different modalities of brain functioning [10,11]. Anatomically, the main hubs of SN are the anterior insula and the dorsal part of anterior cingulate cortex (ACC) [12].

Sedative drugs such as midazolam and propofol were shown to reduce DMN and DMN-CEN connectivity [13-16], while stimulant drugs such as cocaine and modafinil were reported to predominantly enhance CEN and CEN-SN connectivity [17-20]. Under acute GHB challenge with moderate doses, a previous study revealed an acute increase of the SN-DMN and SN-CEN inter-network rsFC, the latter via a regional seed in the mPFC (dorsal nexus) [21]. A selective increase of cerebral perfusion in the right anterior insula and in the ACC was also observed in this study, suggesting a crucial role of SN in mediating the acute effects of GHB administration. To date, no study has investigated the post-acute fMRI signal patterns after nocturnal GHB.

In contrast to the growing knowledge of their functional properties, the neurochemical regulation of large-scale brain networks remains largely unknown. Studies applying combined magnetic resonance spectroscopy (MRS) and fMRI reported significant associations between the spectral signals of main inhibitory and excitatory neurotransmitters, gamma-aminobutyric acid (GABA) and glutamate (Glu), and FC, suggesting their role in synchronizing brain activity across specific regions [22-26]. Importantly, this was demonstrated for both regional and inter-regional (task-dependent) FC, but the direction of the association was variable depending on the brain region considered and on task-demand [26-28]. Moreover, most studies were focused on specific interactions between single network nodes rather than global modulation of network activity [24] and pharmacological studies combining rsFC and neurometabolic investigations in humans are still limited.

To elucidate the neural underpinnings of the above described wake-promoting effects of GHB, we investigated whole-brain rsFC of the DMN, the CEN, and the SN, combined with GABA/Glu ratios in the ACC in the morning after nocturnal application of the drug in 16 healthy male volunteers using a placebo-controlled, double-blind, randomized, cross-over pharmacological fMRI design. The MRS-seed in the ACC was selected as recent studies demonstrated that neurochemical balance in this brain region modulate network interconnectivity [29], and the involvement of the ACC was consistently reported in acute challenges with GHB [21,30-32]. We hypothesized that GHB would switch the rsFC configuration into an extrinsic CEN-directed brain state, coherently with its known post-acute wakefulness-enhancing effects. We expected a significant interaction of altered Glu and GABA levels in the ACC with global changes of rsFC patterns, thus supporting a crucial role of neurochemical balance in this brain area in modulating neural effects by GHB.

## Methods

### Permission

The study was approved by the Swiss Agency for Therapeutic Products (Swissmedic) as well as by the Ethics Committee of the Canton of Zurich and registered at ClinicalTrials.gov (NCT02342366). All participants provided written informed consent according to the declaration of Helsinki.

### Study design

The study followed a randomized, placebo-controlled, order-balanced, double-blind, cross-over design. Two experimental nights (GHB vs. placebo) were separated by a washout phase of seven days. Prior to definitive enrollment into the study, all participants underwent a polysomnographic examination in the sleep laboratory of the Institute of Pharmacology and Toxicology of the University of Zurich to exclude sleep-related disorders such as sleep apnea, restless legs syndrome, sleep onset rapid-eye movement (REM) sleep and reduced sleep efficiency (< 80%). To allow for habituation to the sleep laboratory setting, each experimental night was preceded by an adaptation night. Apart from the here presented post-sleep fMRI RSN results, GHB effects on sleep neurophysiology [31], kynurenine pathway metabolites [33], and post-awakening brain metabolite signals and vigilance [34] were also assessed in the experiment and published elsewhere.

### Participants

Twenty healthy, male volunteers completed the study, whereof 4 participants were excluded from the final data analysis due to technical issues with the MR scanner or insufficient MR data quality (mean age of included participants: 25.8±5.1 years). The following criteria were required for inclusion: (i) male sex (to avoid potential impact of menstrual cycle on primary outcome variables [35]; (ii) age within the range of 18 to 40 years; (iii) absence of somatic, neurologic, and psychiatric disorders; (iv) no first-degree relatives with a history of highly heritable psychiatric disorders such as schizophrenia, bipolar disorder, autism, and attention-deficit/hyperactivity disorder (ADHD); (v) non-smoker; (vi) no history of regular drug use (lifetime use <5 occasions of each drug, except occasional cannabis use). No participant reported previous experiences with GHB in their life. Participants had to refrain from illegal drugs for two weeks and from caffeine for one week prior to the first experimental night and throughout the entire study. No use of alcohol was allowed 24 h before each study night. Participants were instructed to keep a regular sleep-wake rhythm with eight hours time-in-bed from 23:00pm to 07:00am during one week prior to the first experimental night and in the week between the two experimental nights. To ensure compliance with this requirement, participants wore an actimeter on the non-dominant arm and kept a sleep-wake diary. All volunteers received a monetary compensation for their study participation.

### Urine Immunoassay

Urine samples were taken on each test night, to ensure abstinence from illegal drug use (Drug-Screen Multi 12-AE, Nal von Minden GmbH, Regensburg, DE).

### Drug administration

At each experimental night, study participants were awoken at 02:30am to receive 50 mg/kg of GHB (Xyrem^®^) or placebo dissolved in 2 dl of orange juice, matched in appearance and taste (see Figure 1). This dose represents the maximal therapeutic starting dose in narcolepsy. After GHB/placebo intake, volunteers where allowed to immediately return to sleep. GHB administration in the middle of the night was chosen because of its short half-lifetime and its potency to enhance deep sleep in the second half of the night, after the physiological dissipation of sleep intensity and propensity [31].

**Figure 1.**
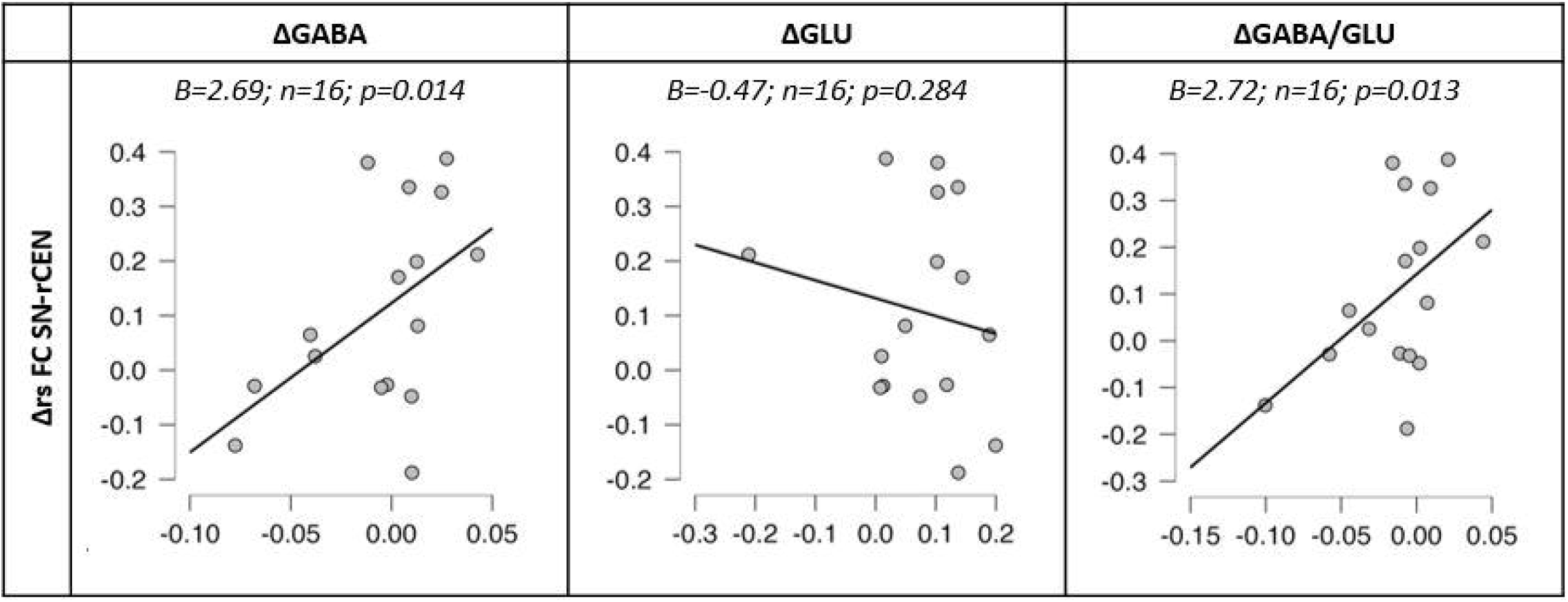
Study design of the experimental nights. Sleep period (23:00-07:00), time point of drug administration (02:30) and MRS/MRI scan (09:00) are indicated.

### MRI data acquisition

The fMRI resting state scan was performed in the morning after both experimental nights on a Philips Achieva 3T whole-body MR-unit equipped with a 32-channel head coil (Philips Medical Systems, Best, The Netherlands). The session started at 09:00am with a T1-weighted anatomical brain scan and was followed by fMRI acquisition (5 and 10 min duration respectively). RsFC time series were acquired with a sensitivity-encoded single-shot echo-planar imaging sequence (SENSE-sshEPI). The rsFC protocol used the following acquisition parameters: TE=35ms, TR=3000ms, flip angle=82°, FOV=220mm, acquisition matrix=80×80 (in plane voxel size=2.75×2.75mm^2^), 32 contiguous axial slices (placed along the anterior-posterior commissure plane) with a slice thickness of 4⍰mm and a SENSE factor R=2.0. For structural reference, a magnetization prepared rapid gradient-echo (MP-RAGE) T1-weighted anatomical scan was acquired with the following parameters: TR/TE=9.3/4.6ms, flip angle=8°, 160 sagittal slices, FOV=240×240mm, voxel size=1×1×1mm.

### MRI data preprocessing

Standard image data preparation and pre-processing as well as statistical analysis and visualization were performed in Matlab (The Mathworks Inc., United States) and BrainVoyager (Brain Innovation B.V., The Netherlands). Functional data preprocessing included a correction for slice scan timing acquisition, a 3D rigid body motion correction, a spatial smoothing (gaussian kernel of 6mm full-width-half-maximum), a temporal high-pass filter with cut-off set to 0.0080Hz per time-course and a temporal low-pass filter (gaussian kernel of 3s). The mean frame wise displacement was estimated from each time-series prior to nuisance and motion regression to exclude motion-driven bias in connectivity correlations [36]. For further details, see supplementary materials and previous publications [21,37].

### Statistical analysis of fMRI images

#### Independent component analysis of resting-state fMRI networks

The ICA analysis of RSN networks followed the identical approach described in a previous study of ours [21]. The analysis is based on a hierarchical approach specifically designed to study FC under changing experimental conditions (see also [38]) in which first-level (single-subject, single-scan) and second-level (group) analyses are performed using the fastICA [39] and the self-organizing group-level ICA (sogICA) algorithm [40]. For each scan condition (GHB vs. placebo), 30 ICA components were extracted using fastICA, roughly corresponding to 1/6 of the number of time points (see also [41] [42]). A more detailed description is available in supplementary materials and previous publications [21].

#### Analysis of correlations between RSN-FC and single voxel MRS metabolites

To explore possible interactions between neurochemical brain balance and RSN-FC, we also investigated associations of FC alterations between conditions with the respective changes of metabolite signals measured by single voxel (sv-) MRS analysis. The sv-MRS was performed at each experimental session immediately before the fMRI paradigm and MRS spectra were acquired from a voxel placed in the anterodorsal portion of the ACC (see MRS-protocol details in a separate publication [34]). Additionally a GABA/Glu ratio was calculated ([GABA/Glu ratio]=[GABA signal]/[Glu signal]) as an index of inhibitory/excitatory equilibrium [29,43,44]. The change in metabolite spectral signals across experimental conditions was calculated subtracting the metabolite signlas of the GHB condition from signals of placebo condition (e.g. [ΔGABA]=[GABA at placebo condition]–[GABA at GHB condition]).

For internetwork connectivity, the change of correlation-z-values (ΔrsFC) across conditions in the internetwork connectivity matrices was calculated subtracting the z-value of GHB condition from z-value of placebo condition ([ΔrsFC]=[z-value at placebo session]–[z-value at GHB session]) for all network-pairs showing significant changes of between-network connectivity in the paired t-test of connectivity matrices (p<0.05). Finally, generalized linear models with normal distribution and identity link function were performed to assess associations between ΔrsFC for significantly altered internetwork connections and metabolites changes expressed as ΔGABA, ΔGlu and ΔGABA/Glu-Ratio, after controlling for experimental session order (GHB first vs. placebo first).

### Subjective state variables

Each participant’s post-awakening mental state was assessed at 10:00am using the self-report questionnaire EWL-60 (“Eigenschaftswörterliste”) [45]. The EWL-60 is an established rating scale assessing multidimensional aspects of subjective mental state, which has been found to be well suited to measure short-term changes induced by psychoactive drugs [46]. It is composed of a list of 60 adjectives (e.g., “active”, “sorrowful”, “tired”, “sociable”), which participants have to rate on four-point Likert scales ranging from 0 (not at all) to 3 (very much). Single item scores can be grouped into six subscales (i.e., performance-related activation, general inactivation, extro-/introversion, general well-being, emotional sensitivity, depressiveness/anxiety). Generalized linear models with normal distribution and identity link function were performed to assess associations between EWL subscale scores and condition (GHB vs. placebo), after controlling for experimental session order. All statistical tests were carried out at a significance level of p<0.05 and were performed using SPSS 26.0, R-Studio and JASP.

## Results

### fMRI data and resting-state networks

For both conditions (GHB and placebo), the mean frame-wise displacement was below the critical threshold of 0.5 mm and did not differ between the conditions. Using the network template masks for extracting the homologue network best-fitting ICA components from each subject, we examined the differences between conditions (GHB vs. placebo), in both within-network (via voxel-wise analysis) and between-network (via correlation analysis) connectivity. No significant drug effects were found within the DMN, the CEN, and the SN. In the internetwork functional connectivity analysis, we observed a significant effect in the internetwork connectivity of the right CEN (rCEN) with the SN in the GHB condition (one sample t-test: p=0.038), which was not present under placebo (one sample t-test: p=0.460). Coherently, rCEN-SN internetwork connectivity was significantly higher in the GHB condition compared to placebo (GHB vs. placebo, paired t-test: p=0.017) (see Figure 2).

**Figure 2.**
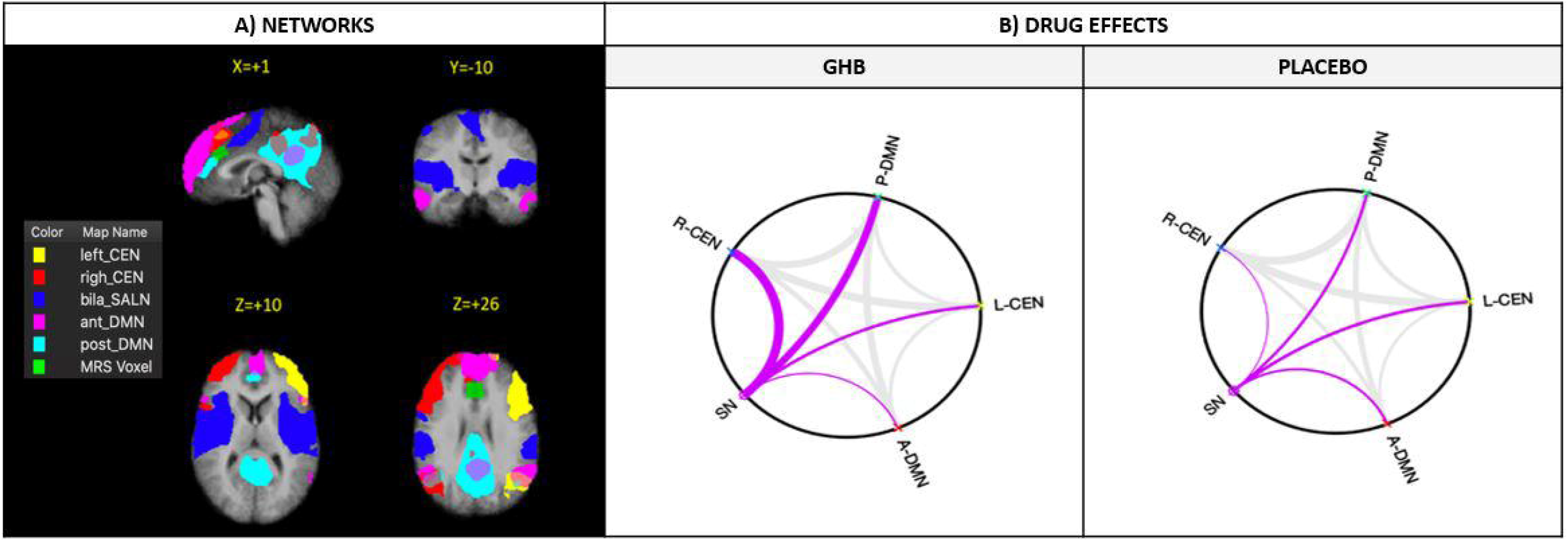
Resting-state networks (RSNs) and internetwork connectivity patterns. (A) All RSNs are shown in three planes on average Talairach anatomical scan. The MRS-voxel is shown in all planes (green). (B) The anterior and posterior default mode network (A-DMN, P-DMN), the left and right central executive network (L-CEN, R-CEN) and the salience network (SN) are considered. The network graph highlights the connections of the SN with the other RSNs (B).

### Associations between RSN-rsFC and sv-MRS metabolite signals

Generalized linear regression models revealed that ΔrsFC of the rCEN-SN was significantly associated with ΔGABA (B=2.69; SE=1.10; 95% CI: 0.54, 4.84; p=0.014) and ΔGABA/Glu-Ratio (B=2.72; SE=1.10; 95% CI: 0.57, 4.88; p=0.013) but not with ΔGlu (B=-0.47; SE=0.44; 95% CI: −1.33, 0.39; p=0.284), after correction for experimental session order (see Figure 3).

**Figure 3.**
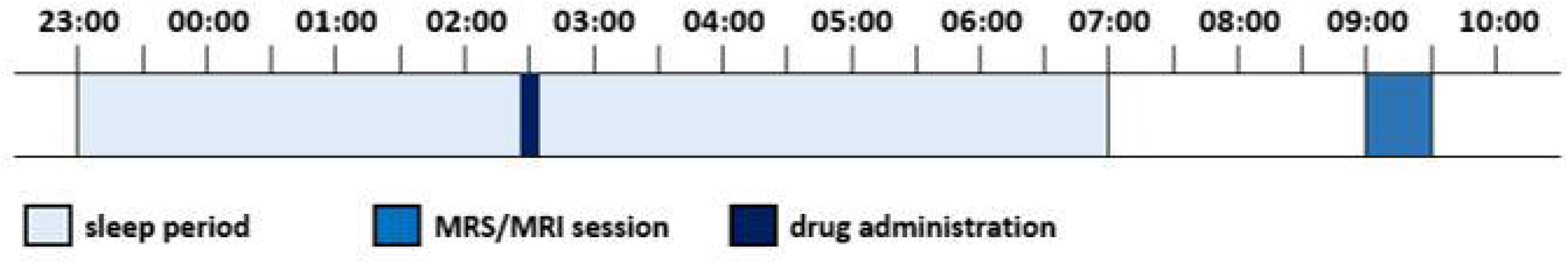
Scatterplots showing associations between the GHB-induced changes in resting-state functional connectivity between the salience network and right central executive network connection (ΔrsFC SN-rCEN) and the GHB-induced changes of metabolite signals in the anterior cingulate cortex (ΔGABA, ΔGlu, ΔGABA/Glu). In the y-axis, the change of rsFC is obtained by extracting the correlation-z-values in the internetwork connectivity matrices. The changes of metabolite signals in the x-axis are reported as mean differences of relative Cramér-Rao lower bounds. Generalized linear models with normal distribution and identity link function were performed.

### Subjective drug effects

Generalized linear regression models revealed no significant effects of condition (GHB vs. placebo) and experimental session order on morning EWL subscale scores (all p>0.13, see supplementary materials).

## Discussion

The present study aimed at investigating the neuropsychopharmacological effects of a nocturnal dose of 50 mg/kg GHB p.o. on next mornings’ rsFC and its relationship to GABA and Glu alterations in the ACC. First, we observed a newly induced rCEN-SN coupling after a night with GHB, which was not present in the placebo condition; second, we found that this rsFC alteration was significantly associated with GABA changes and GABA/Glu ratios in the ACC.

In previous studies, GHB was found to acutely modulate rsFC patterns. In particular, GHB administration (35mg/kg p.o.) in wake healthy individuals acutely increased rsFC between the DMN and the SN (at 34min after GHB-intake) compared to placebo, without significantly affecting the spatial rsFC distribution of all major large-scale networks [21]. Consistently, the here reported post-acute alterations across the same large-scale networks did not affect the within-network rsFC distributions, and the observed switching from DMN-SN (acute, previous studies) [21] to rCEN-SN (post-acute, current study) coupling confirms the SN being a critical target of GHB. In line with this hypothesis, a local increase in cerebral blood flow under acute GHB was also detected in main hubs of the SN, such as the ACC and the right anterior insula, which were correlated with increased relaxation, and body/emotion awareness [30]. Similarly, current source density EEG analysis revealed significant spectral alterations in the ACC under acute challenge with 35mg/kg (wake condition) [32] and 50mg/kg GHB p.o. (sleep condition) in two separate observations [31]. Thus, activation of the SN, and thereby modulation of the balance between DMN and CEN is a core neuronal effect of GHB at both acute and post-acute phases.

These findings are particularly intriguing when considering the detailed functional meaning of the SN and the unique psychopharmacological and behavioral profile of GHB. The SN has been proposed as a detector of saliency to guide adaptive behavior [10]. Anchored in the anterior insula, the dorsal ACC, as well as other subcortical structures, the SN responds to biologically relevant internal and external stimuli and modulates the switch into different brain states by regulation of CEN/DMN activity. Accordingly, SN dysfunctions were frequently associated with different symptom dimensions in neuropsychiatric disorders. While increased rsFC between main SN and DMN hubs was consistently related to depressive symptoms and ruminations [47,48], intrinsic alterations in the SN and disturbed CEN-DMN transition were linked to reality distortion and cognitive deficits in schizophrenia spectrum disorder [11].

Now, translating these findings to GHB neuropsychopharmacology, one may expect to observe different network patterns coherently to the different behavioral effects of GHB in the acute and post-acute phase. In the acute phase, 35 mg/kg oral GHB induced mixed sedative/stimulant behavioral effects, which were accompanied by increased SN-DMN connectivity [21]. This rsFC pattern indicates a switch into a more internally-directed brain state, compatible with reduced attentional focus to external stimuli [1,49-51]. Similar rsFC patterns were also observed during mindfulness, light propofol sedation, and increased homeostatic sleep pressure after sleep deprivation [49,52,53]. On the contrary, the post-acute behavioral effects of nocturnal 50mg/kg oral GHB included activation and vigilance enhancement [6,34,54]. Thus, the increased rCEN-SN rsFC we observed in this study is coherent with the previously reported stimulant effects of the substance and suggests a post-acute switch into a more externally directed state of brain functioning. In line with that notion, increased rsFC between hubs of the CEN and SN was already related to enhanced attentional focus in meditation practioners at rest [55] and to symptoms improvement in children with ADHD treated with methylphenidate [20]. Moreover, reduced rsFC between main hubs of the CEN and SN was frequently associated with fatigue and daytime sleepiness in disorders such as narcolepsy, depression, chronic fatigue syndrome, fibromyalgia and Parkinson disease [56-59]. Thus, increased CEN-SN coupling could represent a neural signature of stimulant effects of GHB and offer a clinical marker for its therapeutic use. Importantly, we did not find any significant effects of GHB on the subjective mental state in this study. However, we observed increased sustained vigilant attention (assessed with a 10min visual psychomotor vigilance test), which was tested in the same experiment and reported in a previous publication [34].

To investigate how the reported modulation of rsFC alterations relate to the neurochemical brain homeostasis, we analyzed previously published data of MRS from an ACC-seed assessed in the same individuals and from the same experiment [34]. The selection of the ACC-seed to study rsFC-regulation was driven by previous evidence showing that the ACC is a crucial target of GHB effects [21,30-32,34] and numerous reports on the ACCs role in controlling global brain function [28,29]. In the current analysis, ΔGABA and ΔGABA/Glu levels but not ΔGlu levels predicted the increased rCEN-SN coupling. These findings, are coherent with other reports in the literature, which described cingulate GABA to be positively associated with DMN deactivation and increased CEN connectivity [23,27,29]. In particular, Levar and colleagues recently demonstrated GABA levels in the ACC to be positively correlated with rsFC in the left and right CEN, while being negatively correlated to rsFC between CEN and DMN [29]. The authors also described a negative association of GABA/Glx (with Glx considered as glutamine + Glu) levels with rsFC in the left insula and left occipital cortex but no correlations between Glx levels and rsFC. Notably, despite the ACC being itself part of the SN, GABA levels in this area were positively correlated with rsFC in the CEN but not in the SN, supporting the view of a neurochemical balance in the ACC to be involved in functional control of distant brain areas. In our study, we also observed a significant positive association of ΔGABA/Glu ratio with ΔrsFC of rCEN-SN. As bidirectional interaction between Glu and GABA are widely reported in the literature, our results further suggest that the neurochemical equilibrium might be critical for internetwork rsFC control, rather than the single metabolite levels [43,44,60,61].

Our study bears several limitations. MRS provides the total metabolite signals in a given ROI, which is why different metabolite pools (e.g. cytoplasmic, vesicular, or extracellular) cannot be differentiated. Thus, it remains unclear if the observed alterations of Glu are related to increased glutamatergic transmission in the ACC or to other indirect metabolic alterations in the region [34]. Obviously, MRS-data also limits our investigation to the ACC and we cannot exclude concomitant neurochemical modulation of rsFC through other pathways. As such, similar neurochemical alterations could also occur in other crucial SN hubs such as the anterior insula. Moreover, the relevance of mesolimbic dopamine levels for SN regulation was already described in the literature and may also contribute to the effects of GHB on rsFC [50,62]. In addition, our study sample consisted solely of young healthy men, to avoid the possible interference of hormonal fluctuations throughout the menstrual cycle on brain metabolite levels [35]. This, taken together with the small sample size and the resulting low statistical power, does not yet allow to generalize our results to the entire healthy population.

In conclusion, the present study provided evidence of persisting internetwork connectivity changes in the morning following a nocturnal therapeutic dose of GHB in humans. The observed alteration in connectivity pattern seem to indicate a modulation of the balancing function of the SN between the DMN and the CEN, towards a more externally oriented brain state, which is in line with GHB’s ability to improve next-day waking functions. We also described a GHB-induced positive interaction of GABA/Glu balance in the ACC with whole-brain connectivity changes. Thus, our findings support the idea of an excitatory/inhibitory equilibrium in the ACC to be actively involved in the modulation of rsFC on a large-scale level. Future research should clarify the generalizability of these findings to other stimulant and/or sedative drugs affecting GABA and Glu homeostasis and further assess correlations with cognitive and behavioral effects in clinical populations.

## Data Availability

All data produced in the present study are available upon reasonable request to the authors

## Funding and disclosure

All authors report no conflicts of interest, financial or otherwise. The study was supported by grants from the Swiss National Science Foundation (SNSF) to HPL (grant # 320030_163439) and the Clinical Research Priority Program *Sleep & Health* of the University of Zurich.

## Acknowledgements

We thank Vinnie Kandra for supporting us with the data acquisition.

## Author Contributions

FB: data analysis and interpretation, drafting the article; FE: data analysis and interpretation; critical revision of the article; DAD: project conceptualization and planning; data collection; NZ: data collection, data analysis and interpretation; BBQ: project conceptualization and planning, critical revision of the article; PS: data collection; HPL: project conceptualization and design, critical revision of the article; ES: project conceptualization and design, critical revision of the article; OGB: project conceptualization and design, data collection, drafting the article.

## Notes

### Competing Interest Statement

The authors have declared no competing interest.

### Clinical Trial

NCT02342366

### Author Declarations

The study was approved by the Swiss Agency for Therapeutic Products (Swissmedic) as well as by the Ethics Committee of the Canton of Zurich.

