## Supplementay Materials for "Subacute changes in brain functional network connectivity after nocturnal sodium oxybate intake are associated with anterior cingulate GABA/glutamate balance"

### **Supplementary Materials - Methods**

***MRI data preprocessing.*** After 3D rigid body correction, a 24-parameter model of head motion was created for each data set with the purpose of regressing out any residual effects of motion from each data set prior to FC estimation. This model incorporates the 6 head motion translation and rotation parameters estimated during 3D rigid body correction, the 6 first order derivatives of the motion parameters, and the 12 corresponding squared items (1). Structural and functional data were co-registered and spatially normalized to the Talairach standard space using a twelve parameter affine transformation. In the course of this procedure, the functional images were resampled to an isometric 3 × 3 x 3 mm^3^ grid covering the entire Talairach box. Nuisance physiological signals from white matter (WM) and cerebro-spinal fluid (CSF) were estimated from each data set by segmenting the WM and the ventricles in the normalized T1 volumes and calculating the average WM and CSF signals from these volumes. Following previous recommendations (2, 3), all 24 motion parameters, together with the WM and CSF signals, were regressed out from each time-course at each voxel.

***Independent component analysis******.*** To select ICA components, we applied the RSN component templates derived from the baseline scans of a previously published dataset (acquired with the same parameters and the same MRI scanner) (4). We selected the DMN, the SN, and both the left and right CEN (l-CEN, r-CEN) as RSNs of interest and each RSN template was used to select one best-fitting RSN component per subject and per scan for the second-level random-effects statistical analysis (4, 5). The RSN with the highest goodness-of-fit score was chosen as the best-fitting component, defined as the difference between the average component score inside the mask minus the average component score outside the mask (5, 6). For each RSN of interest, all selected ICA component maps were entered into the second-level (group-level) statistical analysis. Besides the within-network connectivity changes within a given RSN of interest, the between-network connectivity changes (i.e. between two RSNs) were also assessed from the ICA component time-courses. Thus, the scan- and subject-specific ICA component time-courses were extracted for all individual ICA components selected as SN, DMN, r-CEN and l-CEN and correlation coefficients (Pearson's r) were computed per scan per subject for all possible pairs of ICA component time-courses for both individual scans. The resulting connectivity matrix was used for a second-level (group-level) statistical analysis after Fisher transformation from r to z values. Thereby, paired t-tests were conducted to compare correlation z values between different experimental conditions (GHB vs. placebo). Mean connectivity regression of the within-subject connectivity matrices was also performed, albeit without pre-whitening due to the choice of low-pass filtering the original time-courses (7, 8).

### **Supplementary Materials – Results**

| **Supplementary Table 1. Subjective drug effects.** | | | | | | |
| --- | --- | --- | --- | --- | --- | --- |
| **EWL subscales** | **Mean ± SD Placebo** | **Mean ± SD**  **GHB** | ***Coefficient B*** | ***Standard error*** | ***Wald χ2*** | ***P value*** |
| Performance-related activation | 19.1 ± 4.4 | 19.1 ± 4.8 | 0.07 | 1.59 | 0.002 | 0.96 |
| General inactivation | 15.8 ± 3.7 | 16.8 ± 6.3 | -1.03 | 1.76 | 0.34 | 0.56 |
| Extro-/introversion | 19.8 ± 2.9 | 19.6 ± 3.6 | 0.19 | 1.12 | 0.29 | 0.87 |
| General well-being | 21.3 ± 5.1 | 20.0 ± 6.0 | 1.38 | 1.91 | 0.52 | 0.47 |
| Emotional sensitivity | 15.4 ± 3.5 | 16.0 ± 4.2 | -0.71 | 1.32 | 0.29 | 0.59 |
| Depressiveness/anxiety | 8.4 ± 0.7 | 9.1 ± 2.0 | -0.77 | 0.52 | 2.22 | 0.14 |
| Generalized linear model with normal distribution and identity function. Dependent variable: EWL subscores; factor: treatment (placebo/GHB); co-variable: treatment order. Abbreviations: EWL: Eigenschaftswörterliste; SD: standard deviation. | | | | | | |

**Supplementary Materials – Literature**
